# Gabapentin repurposing for glioblastoma therapy: Real-world data analyses augmented by use of active comparators

**DOI:** 10.1101/2025.09.24.25335799

**Authors:** Christine Ann Pittman Ballard, Kevin M. Goff, Mallika P. Patel, Kyle M. Walsh, Michelle Monje, Quinn T. Ostrom

## Abstract

Glioblastoma ‘hijacks’ neuronal pathways to drive tumor growth, and drugs affecting the function of these pathways may potentiate survival gains. Recent studies have suggested clinical benefits with post-diagnostic use of gabapentin. We assessed the impact of taking gabapentin after glioblastoma diagnosis utilizing an active comparator model in a population-based dataset of older adults in the United States. We leveraged a cohort of glioblastoma patients >65 years old who received resection, radiation, and temozolomide from the Surveillance, Epidemiology and End Results data paired with Medicare claims. Those receiving post-diagnostic gabapentin (TMZ+G) were compared to those receiving standard of care treatment only (TMZ), and two active comparators (duloxetine [TMZ+D], and levetiracetam [TMZ+L]). Association between medication use and overall survival was assessed using cox proportional hazards models adjusted for known prognostic factors. Out of 2,494 individuals, 797 (32%) received TMZ, 146 (5.9%) received TMZ+G, 38 (1.5%) received TMZ+D, and 1,513 (60.7%) received TMZ+L. Median survival among those receiving TMZ (10 months) as compared to all other groups (TMZ+G=16.3 months, TMZ+D=16 months; TMZ+L=13.0 months). TMZ+G was associated with 47% decrease in hazard of death (p<0.001) compared to TMZ, and a 32% decrease (p<0.001) compared to TMZ+L. Women had a 43% decrease in hazard of death (p<0.001) in TMZ+G as compared to TMZ+L, while this difference was non-significant in men (p=0.204). These results show survival benefit associated with gabapentin and supports ongoing work therapeutically targeting neuron-glioma interactions.

---

The development of novel drugs is characterized by immense investments of both money and time, compounded in glioblastoma (GB) by designing blood-brain barrier (BBB)-penetrant compounds. Drug repositioning (*i.e*., drug repurposing) is a method of finding new uses for existing therapies with known safety and pharmacokinetic profiles, allowing faster and less expensive translation into the clinic. Gliomas ‘hijack’ neuronal pathways to drive tumor growth, and preclinical evidence suggests that drugs affecting neuron-glioma interactions may influence survival gains.^1-9^ Studies by us and others have identified candidate drugs that affect neural signaling (*e.g*. fluoxetine^10^, beta-blockers^11^, and gabapentin^12^) and are associated with extended GB patient survival in real-world datasets. Here, we use an active comparator intention-to-treat model to assess survival differences associated with gabapentin use following GB diagnosis in a population-based dataset of older U.S. adults.

We leveraged the NCI’s Surveillance, Epidemiology and End Results (SEER) data linked to traditional Medicare claims to investigate gabapentin’s effect on overall survival in patients with GB (International Classification of Diseases, Oncology 3^rd^ edition morphology codes 9440/3-9442/3, and topography codes C71.0-C71.9) receiving resection (biopsy-only cases excluded), radiation (≥2 claims), and temozolomide, and diagnosed from 2008-2019 (followed through 2020). Patients were excluded if <66 years at diagnosis, not enrolled in Medicare Parts A/B/D for equal time, missing follow-up, or diagnosed by autopsy/death certificate. *IDH1/2* mutation was not available for most patients; but among those with available data (2018-2019), all were IDHwt. Presence of seizures and performance status were not available. In addition to a standard-of-care comparator, active comparators or drugs used for a similar indication with a different mechanism of action were used to adjust for potential confounding due to comorbidity status and immortal time bias (*e.g*. bias created by individuals required to survive to reach a specific event after index date^13^) Levetiracetam was selected as an active comparator for gabapentin because both are prescribed for seizure control in glioblastoma patients, while duloxetine was selected as both are prescribed for pain control. We utilized an intent-to-treat protocol (≥2 claims considered “on treatment”) and the first filled treatment was considered for the analysis. Indication for drug use was not known. Associations were assessed using cox proportional hazards models adjusted for age at diagnosis, sex, race/ethnicity, Charlson comorbidity score (six months prior to diagnosis), and extent of resection.

There were 2,494 individuals (47% female; 88% Non-Hispanic White) available for analysis, all of whom received resection, radiation and temozolomide. Of these, 797 (32%) received temozolomide alone (TMZ) and 146 (5.9%) received gabapentin/temozolomide (TMZ+G). Among those receiving selected active comparators, 38 (1.5%) received duloxetine/temozolomide (TMZ+D), and 1,513 (60.7%) received levetiracetam/temozolomide (TMZ+L). Median survival was lowest for TMZ (10 months, 95%CI=9.14-10.9) as compared to all other groups (TMZ+G: 16.3 months, TMZ+D: 16 months; TMZ+L: 13.0 months). In the adjusted model comparing to TMZ alone, TMZ+G was associated with a 47% decrease in hazard of death (HR=0.53, p<0.001), while TMZ+D had a 38% decrease (HR=0.62, p<0.001), and TMZ+L had a 23% decrease (HR=0.77, p<0.001). TMZ+G had a 32% decrease in hazard of death (HR=0.68, p<0.001) compared to TMZ+L only, but no significant differences from TMZ+D (p= 0.352). Women had a 43% decrease in hazard of death (HR=0.57, p<0.001) in TMZ+G as compared to TMZ+L, while this difference was non-significant in men (HR=0.85; p=0.204). There was significant statistical interaction between female sex and TMZ+G compared to TMZ+L (p=0.035) and TMZ alone (p= 0.082). Sex-specific comparisons to TMZ+D were not possible due to small sample size.

These results show survival benefit associated with gabapentin use as compared to TMZ alone and active comparator group TMZ+L and supports ongoing work suggesting that neuron-glioma interactions represent viable therapeutic targets. While this shows a suggestive survival improvement association with duloxetine, estimates are unstable and were not able to be fully evaluated due to small sample size. A female sex advantage in GB survival has been well documented, but sex differences in the pharmacokinetics of duloxetine, levetiracetam, and gabapentin has been studied with variable results.^14^

While valuable for hypothesis generation, retrospective observational studies have multiple significant sources of bias for evaluation of drug efficacy, including lack of randomization leading to unbalanced groups and inflated survival in the treated group due to immortal time bias. Indication for use of drug is not known, and variations in comorbidities or symptoms between comparison groups could create bias. There are also several important limitations to use of Medicare data, which we have discussed previously.^11^ For providers, retrospective analyses can impact prescribing patterns, especially in diseases with limited treatment options such as GB. Previous reports of survival benefit associated with use of neuro-active drugs has received significant attention, leading patients to request off-label use of these therapies into their treatment plan. Rapid evaluation of causal survival associations for these novel potential therapies is needed to better contextualize research results for patients and providers.

## Data Availability

All data are provided under a data use agreement with SEER-Medicare and can be requested at https://healthcaredelivery.cancer.gov/seermedicare/

## Acknowledgments

This study used the linked SEER-Medicare database. The interpretation and reporting of these data are the sole responsibility of the authors. The authors acknowledge the efforts of the National Cancer Institute; Information Management Services (IMS), Inc.; and the Surveillance, Epidemiology, and End Results (SEER) Program tumor registries in the creation of the SEER-Medicare database. The collection of cancer incidence data used in this study was supported by the California Department of Public Health pursuant to California Health and Safety Code Section 103885; Centers for Disease Control and Prevention’s (CDC) National Program of Cancer Registries, under cooperative agreement 1NU58DP007156; the National Cancer Institute’s Surveillance, Epidemiology and End Results Program under contract HHSN261201800032I awarded to the University of California, San Francisco, contract HHSN261201800015I awarded to the University of Southern California, and contract HHSN261201800009I awarded to the Public Health Institute. The ideas and opinions expressed herein are those of the author(s) and do not necessarily reflect the opinions of the State of California, Department of Public Health, the National Cancer Institute, and the Centers for Disease Control and Prevention or their Contractors and Subcontractors.

## Figure Key

**Figure 1.**
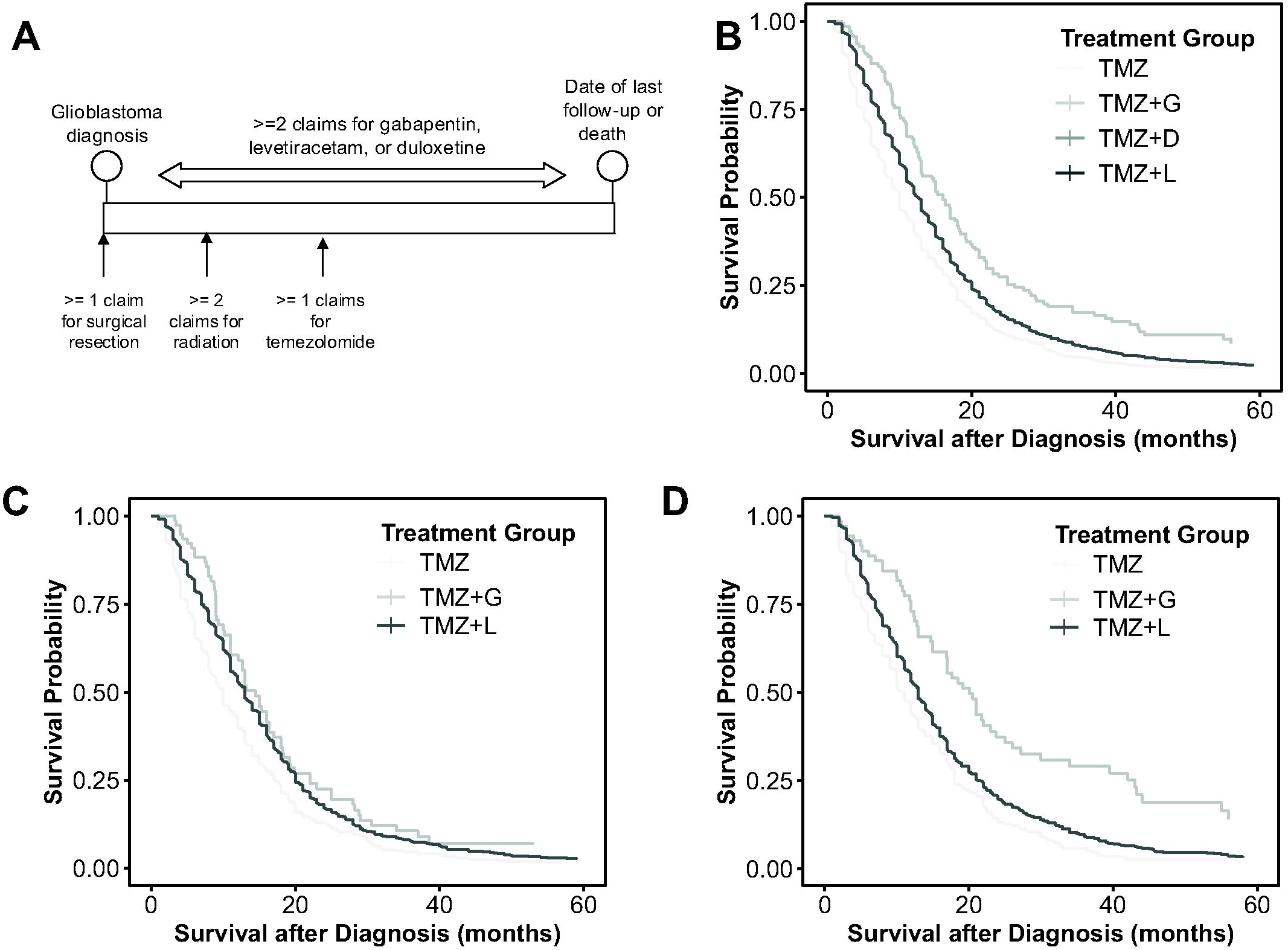
A) Timeline of study design, B) adjusted survival probability in the overall model, C) adjusted survival probability in males only, and D) adjusted survival probability in females only.

